# A national landscaping survey of critical care services in hospitals accredited for training in a lower-middle income country: Pakistan

**DOI:** 10.1101/2020.04.22.20071555

**Authors:** Madiha Hashmi, Arshad Taqi, Muhammad Iqbal Memon, Syed Muneeb Ali, Saleh Khaskheli, Muhammad Sheharyar, Muhammad Hayat, Mohiuddin Shiekh, Chamira Kodippily, Dilanthi Gamage, Arjen M Dondorp, Rashan Haniffa, Abi Beane

## Abstract

**Purpose:** To describe the extent and variation of critical care services in Pakistan

**Materials and methods:** A cross-sectional survey was conducted in all CCUs recognised for postgraduate training to determine administration, infrastructure, equipment, staffing, and training.

**Results:** There were 220 CCUs registered for training, providing 2166 CCU beds and 1473 ventilators. Regional distribution of CCU beds per 100,000 population ranged from 1.0 in Sindh to none in Gilgit Baltistan (median 0.7). A senior clinician trained in critical care was available in 19 (12.1%) of units, giving a ratio of one trained intensivist for every 82 CCU beds and 0.009 trained intensivists per 100,000 population. One to one nurse to bed ratio during the day was available in 84 (53.5%) of units, dropping to 75 (47.8%) at night. Availability of 1:1 nursing also varied between provinces, ranging from 56.5% in Punjab compared to 0% in Azad Jamu Kashmir. All CCUs had basic infrastructure (electricity, running water, piped oxygen) and basic equipment (electronic monitoring and infusion pumps).

**Conclusion:** Pakistan, a lower middle-income country has an established network of critical care facilities with access to basic equipment, but inequalities in its distribution. Investment in critical care training for doctors and nurses is needed.

## Background

Demand for critical care services continue to grow internationally. Resources remain limited, most notably in low and lower-middle income countries (LMICs). In South Asia, overall improved public health and primary healthcare services in the region, the growing burden of noncommunicable disease, and with it a demand for surgical and trauma care has resulted in a shift in health systems priorities [1,2]. There is thus an increasing demand for critical care services, and the associated manpower, infrastructure and equipment requirements in LMICs.

Understanding the landscape of existing infrastructure, equipment and staffing both between and within countries provides valuable information for those seeking to strengthen critical care services and inform disaster and pandemic planning. Furthermore, mapping critical care services to the clinical characteristics of the patient it serves is a fundamental step in evaluating quality of existing service provision and to identify priorities for research and quality improvement.

Sri Lanka was the first country in South Asia to undertake a comprehensive national survey of critical care services [3]. Since then, regional efforts to map critical care services in Asia have contributed valuable information regarding critical care unit (CCU) bed availability in the region [9]. However, information regarding skills, training and organisational processes (essential to developing strategies for improving the quality of care) remains absent. The Pakistan Registry of Intensive CarE (PRICE) [3], a cloud-based surveillance platform, currently supports a network of 43 CCUs in Pakistan recording over 2000 monthly critical care admissions. PRICE provides near real-time reporting on the epidemiology, severity of illness, treatment, microbiology and outcomes of CCU patients, alongside information regarding work force, unit occupancy, unit acuity, and resource utilisation. This information is used to drive local service evaluation and quality improvement interventions. PRICE is a founding member of the recently established Wellcome-MORU-CRIT Care Asia (CCA).

This paper details a national survey of critical care services in Pakistan including organisational structures, equipment, infrastructure and training capacity.

## Setting

Pakistan consists of four provinces (Balochistan, Khyber Pakhtunkhwa, Punjab, and Sindh), two autonomous territories (Azad Jammu Kashmir, Gilgit-Baltistan) and one federal territory (Islamabad Capital Territory) [4]. Islamabad was included in the province of Punjab for the purposes of this study.

## Methods

A CCU was defined as a clinical area (excluding operating theatres) which had the ability to provide organ support for in-patients, including mechanical ventilation. All hospitals recognised by the Pakistan Medical and Dental Council (PMDC) for internship training and the College of Physicians and Surgeons Pakistan (CPSP) for postgraduate residency training in anaesthesia, internal medicine, general surgery, cardiac surgery, pulmonology, nephrology, cardiology and critical care medicine were contacted by telephone by MH. All such hospitals were invited to participate in the survey if they reported the presence of at least one CCU. Eligible hospitals were asked for the number of CCUs, number of ventilators and asked to nominate a senior CCU doctor or sister in charge to respond to the survey questions. If a nominated contact was unavailable, at least one follow-up call was made for each CCU. The surveys were administered by telephone or online between February 2017 and December 2018. All responses were included in the analysis. The survey instrument including characteristics and organizational structure, infrastructure and human resources was based on the tool pioneered in South Asia by our group [5]. Population per region was obtained from the government census up to January 2018 from publicly available sources [4]. CCUs were defined as open, closed or semi-closed [6].

## Results

One hundred and fifty-one hospitals were identified, of which 30 did not have a CCU and were therefore excluded. All 121 eligible hospitals participated in reporting bed and ventilatory capacity. Two hundred and twenty CCUs were identified in these hospitals providing 2166 critical beds and 1473 ventilators. Of these 220 CCUs, 157 (71.4%), completed the full survey of organisational structure, infrastructure, equipment and human resources. The designated contact was not reachable in 60 CCUs and a further 3 were not available for interview (Figure 1). Table 1 summarizes the main characteristics of the hospitals surveyed. The density and distribution of CCU beds within teaching institutions (total and per 100 000 population) by administrative regions is described in Table 2 and Figure 2. Figure 3 reports population density as a reference.

**Table 1:**
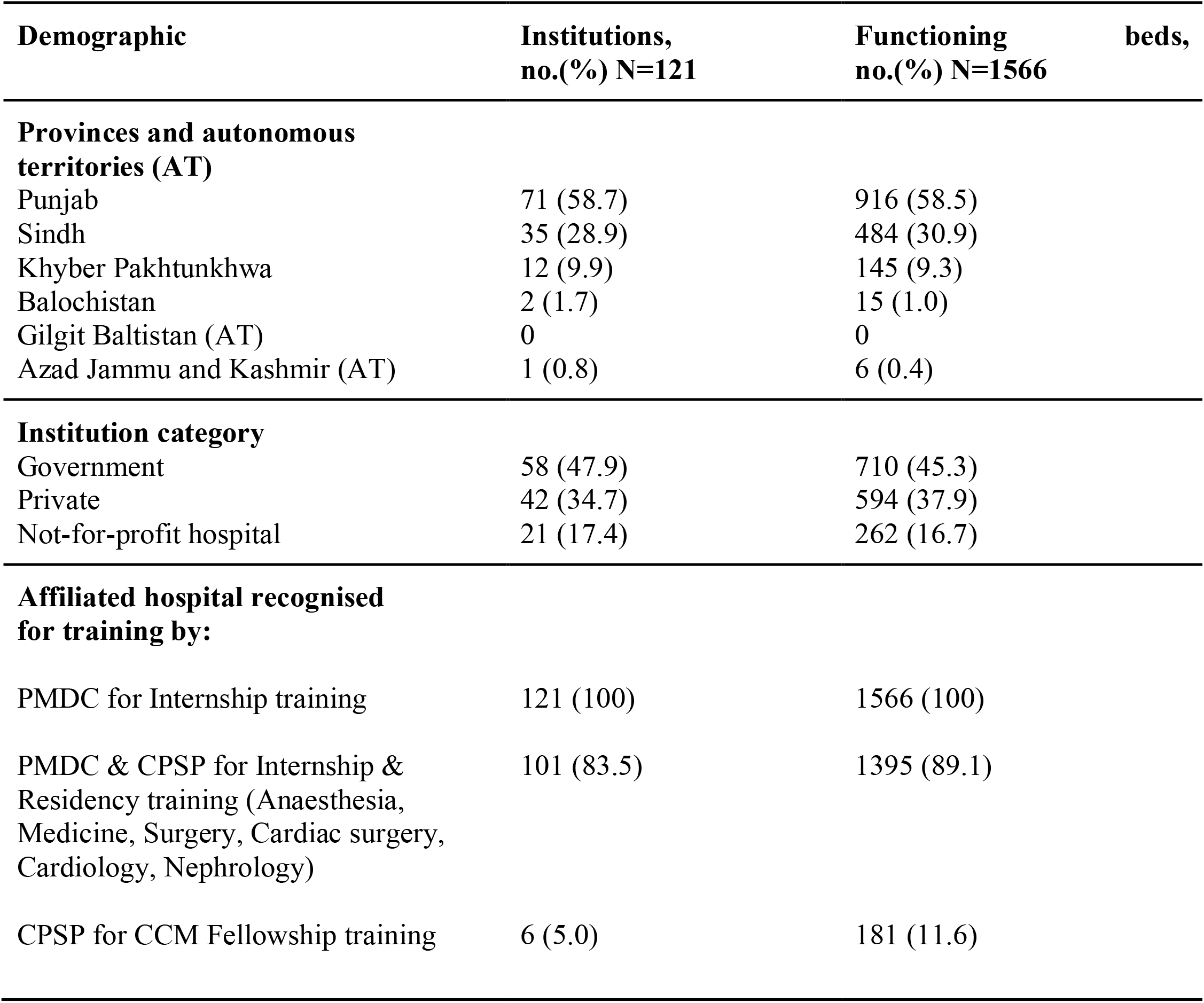
Institutional location and profile.

**Table 2:** Institutions and bed availability per 100 000 population.

| **Administrative Regions** | **Population as per 2017**<br>**census survey** | **Functioning beds per**<br>**100,000 population (total)** |
| --- | --- | --- |
| Pakistan (all 4 provinces and 2<br>independent territories) | 221,613,314 | 0.71 |
| Punjab | 120,012,442 | 0.76 |
| Sindh | 47,886,051 | 1.01 |
| Khyber Pakhtunkhwa | 35,525,047 | 0.41 |
| Balochistan | 12,344,408 | 0.12 |
| Gilgit Baltistan | 1,800,000 | 0 |
| Azad Jammu and Kashmir | 4,045,366 | 0.14 |

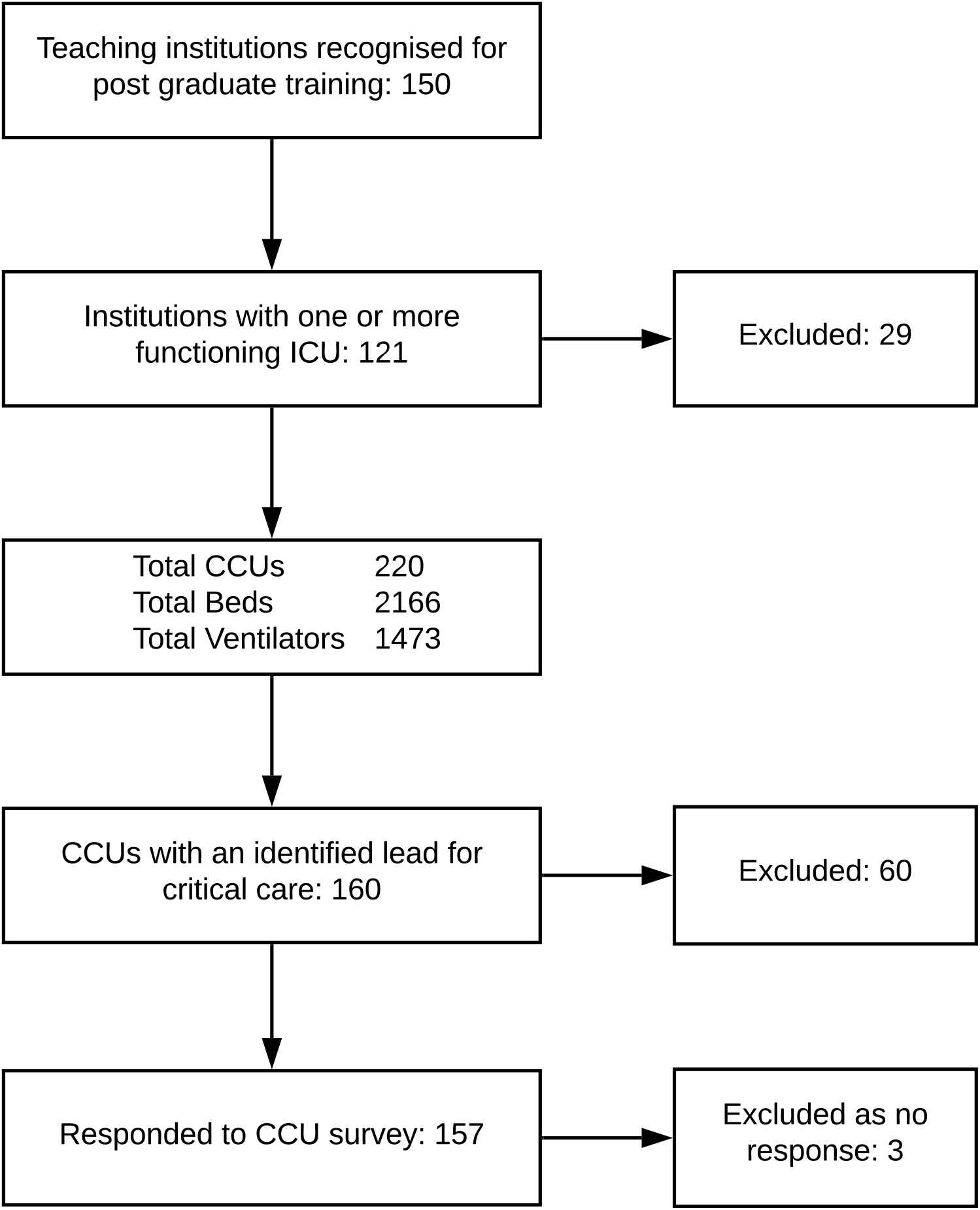

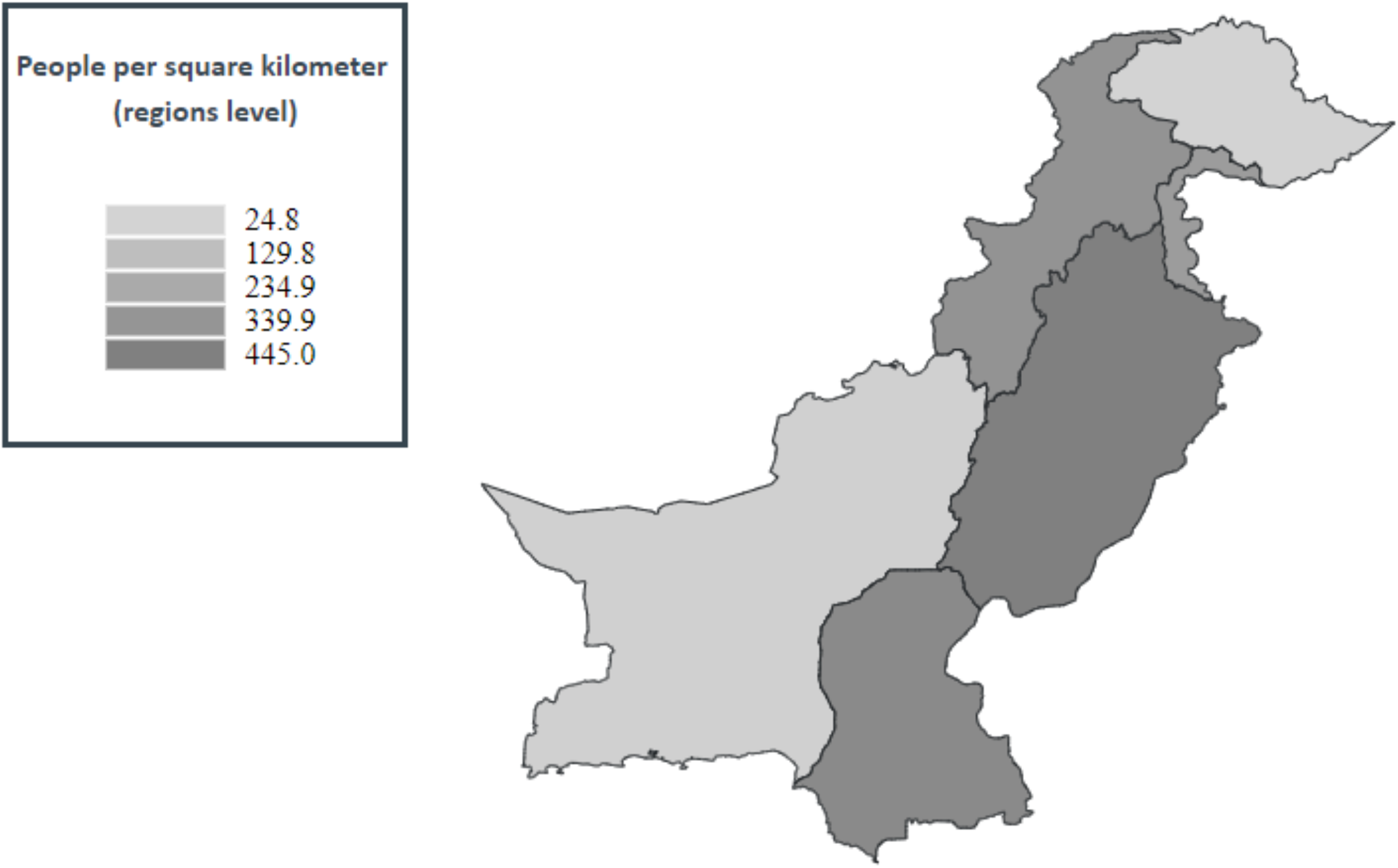

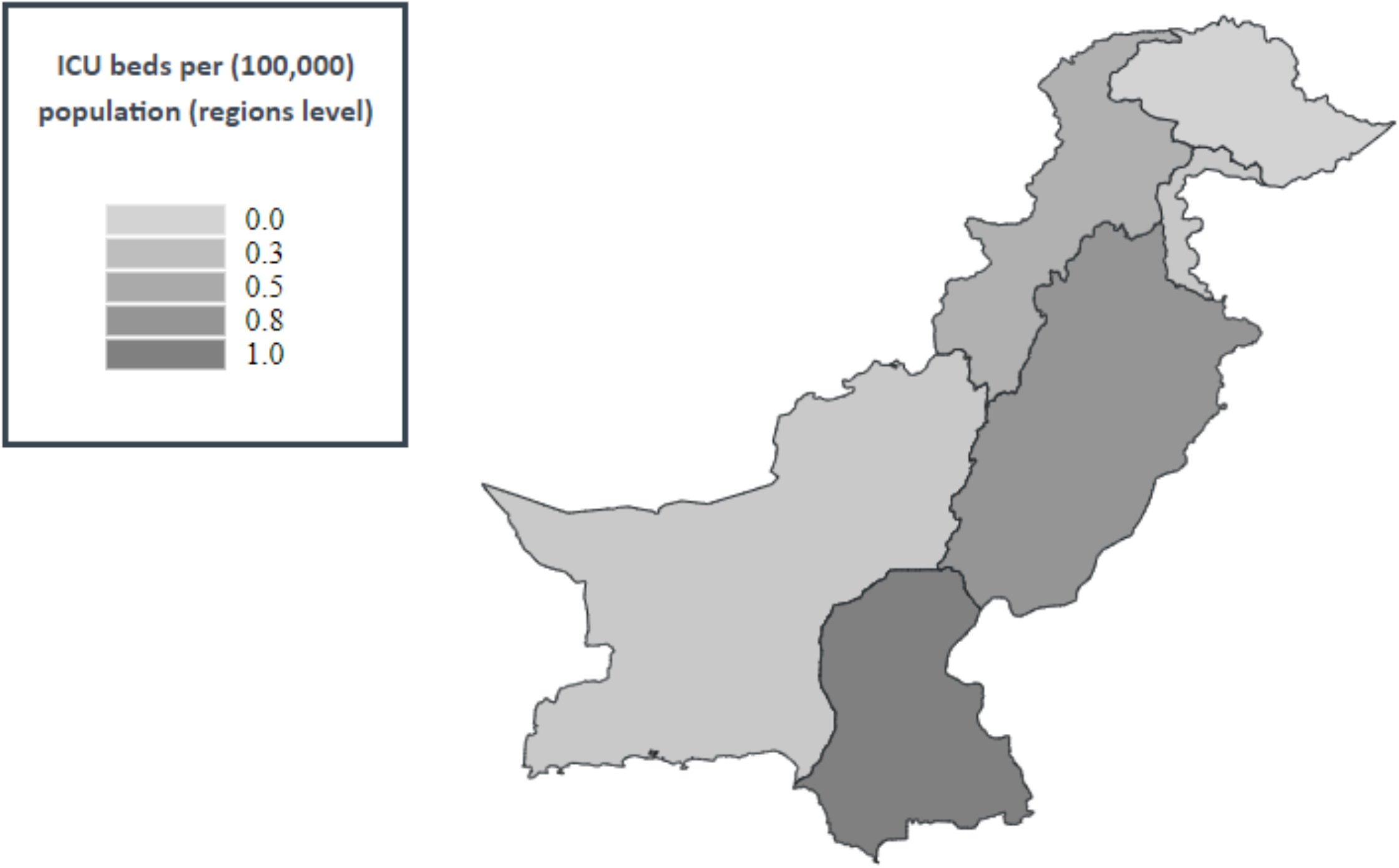

### Access and organisational structure

Average beds within teaching institutions per 100,000 population was 0.7 (total number of CCU beds in participating institutions divided by the total population of Pakistan), ranging from 0 in Gilgit Baltistan to 1.0 in Sindh. The median number of critical care beds per unit was 9 (range 0-30). A total of 58 (47.9%) CCUs were managed directly by the government and 42 (34.7%) of units were managed by the private sector, with the remainder being administered by not-for-profit organisations (17.4%). Seventeen (10.8%) of the CCUs surveyed reported a ‘closed’ model of care whereby intensivists were the consultant in charge of care. Unrestricted visiting for families was practiced in 40 (25.5%) of units (Table 3).

**Table 3:** Critical care unit profile

| **Demographic** | **CCUs, N=157** | **CCU beds, N=1566** | **Ventilators, N=1175** |
| --- | --- | --- | --- |
| **Type of CCU** |  |  |  |
| Medical | 34 (21.7) | 390 (24.9) | 257 (21.9) |
| Surgical | 51 (32.5) | 443 (28.3) | 375 (31.9) |
| General/Mixed | 56 (35.7) | 556 (35.5) | 415 (35.3) |
| Specialized | 16 (10.2) | 177 (11.3) | 131 (11.1) |
| **Model of care** |  |  |  |
| Open | 100 (63.7) | 994 (63.5) | 706 (60.1) |
| Close | 17 (10.8) | 204 (13.0) | 169 (14.4) |
| Semi-closed | 40 (25.5) | 368 (23.5) | 300 (25.5) |

### Infrastructure

Ventilator to bed ratio of 1:1 was observed in 82 (52.2%) of the CCUs (table 4), with Punjab province having the greatest number 49 (59.8%) and Azad Jamu Kashmir the lowest 0. All CCUs had a telephone line, however, only 52.2% had access to the internet. Table 5 summarises the availability of equipment to monitor critically ill patients. Almost all CCUs (95.5%) had access to 1:1 non-invasive multiparameter monitoring. Invasive arterial monitoring and capnography was available in 69 (44.0%) and 9 (5.7%) CCUs respectively. In addition, 51 (32.5%) and 39 (24.8%) units had access to point-of-care haemoglobin and lactate measurement respectively. Isolation rooms essential for management of infectious diseases, including severe acute respiratory infections, were available in 45 (28.7%) CCUs.

**Table 4:** Available infrastructure for each CCU

| **Facilities** | **CCUs, no. (%)** |
| --- | --- |
| Piped oxygen | 154 (98.1) |
| Wall suction units | 132 (84.1) |
| Piped medical air | 147 (93.6) |
| 1:1 bed:ventilator | 82 (52.2) |
| Paediatric mode in all ventilators | 118 (75.2) |
| Syringe pumps | 132 (84.1) |
| Infusion pumps | 125 (79.6) |
| Difficult Airway Trolley (DAT) | 115 (73.2) |
| **Backup automatic electricity generator** |  |
| Yes | 157 (100) |
| **Hand washing facilities in the CCU** |  |
| Yes | 111 (70.7) |
| **CCU isolation rooms** |  |
| Yes | 45 (28.7) |
| **Access to arterial blood gas (ABG) analysis** |  |
| Yes | 142 (90.4) |
| **Location of ABG machine (n=142)** |  |
| Hospital lab | 70 (49.3) |
| Within CCU | 52 (36.6) |
| Another CCU or Operating theatre | 20 (14.1) |
| CCUs reported a POC ABG service.* | 72 |
| **Access to external internet** |  |
| Yes | 82 (52.2) |
| **Telephone** |  |
| Yes | 157 (100) |
| *ABG machines located in the CCU or OR are considered point of care (POC) services. |  |

**Table 5:** Available monitoring facilities

| Facilities | CCUs, no. (%) |
| --- | --- |
| Non-invasive monitoring (including manual central pressure monitoring) | 157 (100) |
| Invasive arterial blood pressure | 110 (70.1) |
| Capnogram | 63 (40.1) |
| Cardiac output (haemodynamics) | 14 (8.9) |
| Point-of-care haemoglobin measurement | 51 (32.5%) |
| Point-of-care lactate measurement | 39 (24.8%) |
| . |  |

### Human resources, team structure and training opportunities

Table 6 summarises the human resources, team structure and training opportunities. A senior clinician trained in critical care was available in only 19 (12.1%) of units. This is a ratio of one trained intensivist for every 82 CCU beds and 0.009 trained intensivists per 100,000 population. The majority of units (86, 54.8) was overseen by a consultant anaesthetist. In the remainder, 38.8% were overseen by a consultant physician and 6.4% by a consultant surgeon. A non-consultant doctor was assigned to CCU round-the-clock with no other work commitments in 140 (89.1%) of CCUs. Of the 121 institutions surveyed, 101 (83.5%) were recognised by the College of Physicians and Surgeons for speciality training (residency training). Critical care medicine (CCM) fellowship training was offered by 6 (4.9%) of institutions.

**Table 6:**
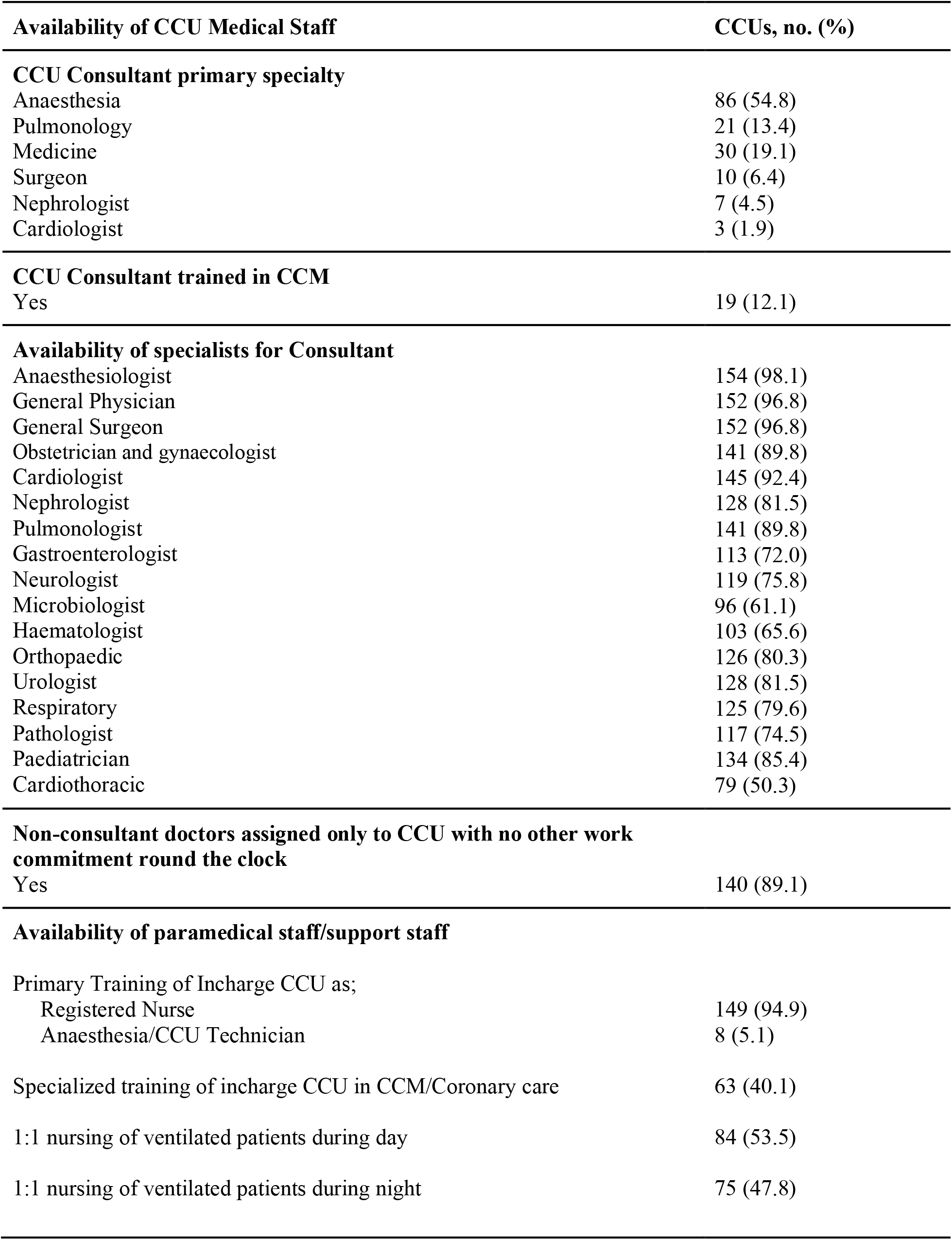
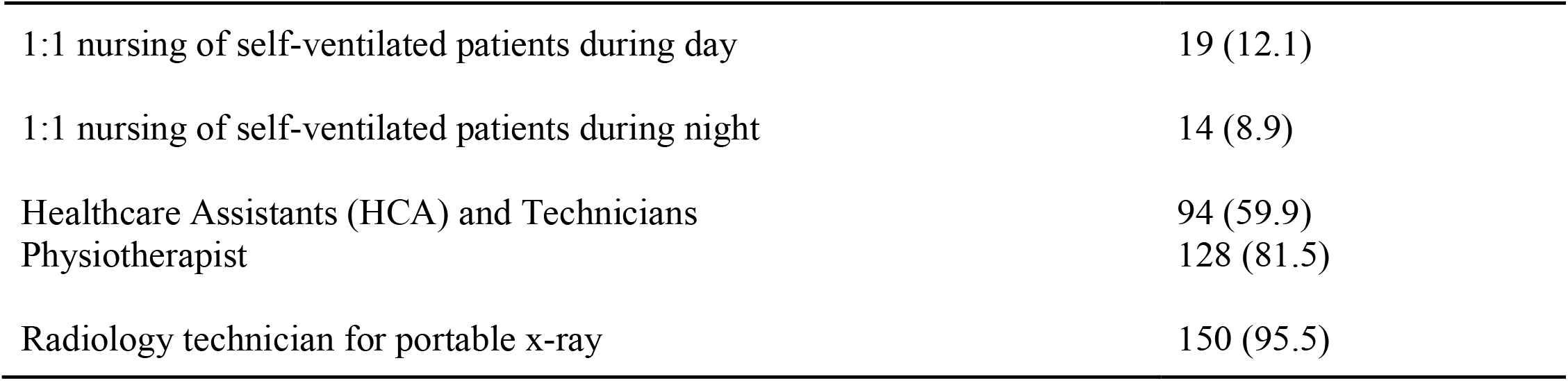
CCU Medical Staffing

| **Availability of CCU Medical Staff** | **CCUs, no. (%)** |
| --- | --- |
| **CCU Consultant primary specialty** |  |
| Anaesthesia | 86 (54.8) |
| Pulmonology | 21 (13.4) |
| Medicine | 30 (19.1) |
| Surgeon | 10 (6.4) |
| Nephrologist | 7 (4.5) |
| Cardiologist | 3 (1.9) |
| **CCU Consultant trained in CCM** |  |
| Yes | 19 (12.1) |
| **Availability of specialists for Consultant** |  |
| Anaesthesiologist | 154 (98.1) |
| General Physician | 152 (96.8) |
| General Surgeon | 152 (96.8) |
| Obstetrician and gynaecologist | 141 (89.8) |
| Cardiologist | 145 (92.4) |
| Nephrologist | 128 (81.5) |
| Pulmonologist | 141 (89.8) |
| Gastroenterologist | 113 (72.0) |
| Neurologist | 119 (75.8) |
| Microbiologist | 96 (61.1) |
| Haematologist | 103 (65.6) |
| Orthopaedic | 126 (80.3) |
| Urologist | 128 (81.5) |
| Respiratory | 125 (79.6) |
| Pathologist | 117 (74.5) |
| Paediatrician | 134 (85.4) |
| Cardiothoracic | 79 (50.3) |
| **Non-consultant doctors assigned only to CCU with no other work commitment round the clock** |  |
| Yes | 140 (89.1) |
| **Availability of paramedical staff/support staff** |  |
| Primary Training of Incharge CCU as; |  |
| Registered Nurse | 149 (94.9) |
| Anaesthesia/CCU Technician | 8 (5.1) |
| Specialized training of incharge CCU in CCM/Coronary care | 63 (40.1) |
| 1:1 nursing of ventilated patients during day | 84 (53.5) |
| 1:1 nursing of ventilated patients during night | 75 (47.8) |
| 1:1 nursing of self-ventilated patients during day | 19 (12.1) |
| 1:1 nursing of self-ventilated patients during night | 14 (8.9) |
| Healthcare Assistants (HCA) and Technicians | 94 (59.9) |
| Physiotherapist | 128 (81.5) |
| Radiology technician for portable x-ray | 150 (95.5) |

The majority of critical care units were managed by registered nurses with general training 149 (94.9%), with the remaining 8 (5.1%) being managed by technicians trained in anaesthesia or critical care. One to one nurse to bed ratio during the day for ventilated patients was available in 84 (53.5%) of units, and in 19 (12.1%) of units for self-ventilated patients. At night this availability dropped to 75 (47.8%) and 14 (8.9%) respectively. Availability of 1:1 nursing also varied between provinces, ranging from 56.5% having a 1:1 availability in the Punjab compared to 0% in Azad Jamu Kashmir. Microbiologists and haematologists were accessible in 96 (61.1%) and 103 (65.6%) of units respectively. Health care assistants or trained technicians were part of the care provision team in 94 (59.9%) of CCUs. Radiology technicians were available in 150 (95.5%) units and a further 128 (81.5%) CCUs had access to physiotherapy services.

## Discussion

This national survey from Pakistan reports very limited critical care bed availability but where available CCUs are well resourced with basic equipment for invasive ventilation and monitoring. It further highlights the lack of critical care trained staff and the need for urgent investment in critical care services to address this gap in training capacity if care is to be improved.

### Critical care capacity

The number of critical care beds in LMICs are known to be lower when compared to high-income countries [7,8], this disparity is pronounced in Pakistan in comparison to neighbouring countries: At 0.71 per 100 000 population, it is lower than Sri Lanka (2.3 critical care beds per 100,000), Nepal (2.8) and India (2.3) [9]. The survey further identified a wide disparity in access to critical care beds between the provinces (Figure 3). Punjab, whilst being the most densely populated province of the country, has lower availability of critical care beds than neighbouring Sindh. Similar disparity exists between major cities in each province (Table 2). As urbanisation and migration to cities for employment continues in Pakistan, and as the burden of non-communicable disease rises - including road traffic accidents and multimorbidities, it is a national priority to address the disparity in access to critical care services [2].

CCUs in both public and private sector institutions (including not-for-profit) had the basic **infrastructure** (electricity and a backup generator, piped oxygen, medical air and suction, infusion and syringe pumps), and basic monitoring (non-invasive multiparameter monitor, mercury thermometer, and manual CVP measurement). Overall ventilator to bed ratio was 1:1.3, meaning 3 out of every 4 CCU beds have the facility to mechanically ventilate. Availability of these resources is reassuring, and suggests that the provision of the mainstays of critical care organ support- ventilation therapy, basic cardiovascular monitoring and support, and delivery of fluids is possible. The safe and effective delivery of these therapies, however, relies not only on the availability of equipment, but on specialist trained staff with the skills to instigate, titrate and troubleshoot treatment.

In contrast to the specific resources of critical care described above, sinks for hand washing were absent in 29.3% of CCUs and access to isolation rooms or cubicles to control cross infection with negative/positive air exchange mechanism was available in just 28.7% of CCUs, the majority of which were private sector tertiary care hospitals in the major cities. Addressing the absence of facilities for infection control is perhaps a key priority for those seeking to improve critical care services in the country, given the increasingly important role critical care plays in the preparation and management of seasonal epidemics (including severe acute respiratory infections-SARI) and in the rising burden of drug resistant infections. Furthermore, access to point-of-care (POC) measurements including lactate haemoglobin, and availability of invasive haemodynamic monitoring, which are increasingly seen as essential resources for the management of critical illness, is lacking [10,11]. Only 39 (24.8%) of units had access to POC lactate and just 14 (8.9%) could invasively monitor haemodynamics. As Pakistan seeks to improve diagnosis and management of critically ill patients with SARI, sepsis and following trauma, better access to POC services and invasive monitoring, along with specially trained staff to interpret and respond to this information, is essential.

### Capacity for training in critical care

The ratio of **trained intensivists** for each CCU bed in this survey is one to 82, much lower than estimates from South Asia, Latin and North America [12–14]. Critical care has been a recognised speciality in Pakistan with a structured training programme since 2004, however, at the time of this survey, only six institutions out of 121 teaching institutions (excluding military sites) were recognized by the College of Physicians and Surgeons of Pakistan for Critical Care Medicine training [15]. To date, just twenty-seven fellows have obtained the fellowship from the College in Critical Care Medicine [16]. Many currently practising intensivists in CCUs still have to travel outside of Pakistan for their higher training fellowships. Low intensivist to patient ratios (< 1:14) in academic medical CCUs have been cited as a barrier to delivery of quality of care and having a detrimental effect on staff well-being, specifically to the quality of pastoral and professional mentorship available for rotating trainees, who may consider specialising in CCM [17]. Lack of specialist training opportunities may be perpetuating the low numbers of designated critical care doctors on-call in CCUs and the low percentage of CCUs which are led by a trained intensivist. Lack of training opportunities extends beyond doctors, with only 40.1% of nurses in charge of CCUs having received any formal training in intensive, critical or cardiac care. Access to microbiologists, specialists who are increasingly considered fundamental to the interdisciplinary management of critically ill patients, was limited (61.1%).

Whilst there is growing evidence to support that intensivist-led patient management is associated with better patient outcomes and greater compliance with broadly accepted indicators of critical care quality [18], a closed model was uncommon in the CCUs surveyed (10.8%). A further 25.5% of CCUs reported a semi-closed structure, but without a trained intensivist as their clinical lead. Investment in the reorganisation of critical care services to improve operational efficiency and patient outcomes (length of stay, duration of mechanical ventilation) in CCUs has resulted in a shift toward closed organisational structures whereby admission to and management of patients within the CCU is coordinated by designated critical care clinicians [18]. Given the paucity of critical care services in Pakistan, such a model of management may promote effective resource utilisation. However, such models require national level investment in specialist training, and institution level investment in hiring and retaining such a specialist workforce. Other settings have demonstrated how investment in critical care trained clinician staff to lead units and investment in training for nurses working in critical care has positively impacted on safety within CCUs and outcomes for critical care patients. Without this investment, efforts to strengthen specialist capacity and improve quality of critical care services through research and implementation will be hindered.

### Limitations

This survey only approached institutions recognised for specialist teaching. Consequently. the number of CCU beds per 100,000 population is underestimated. A recent multicountry snapshot of critical care bed availability [9], to which Pakistan contributed, reported a national average in Pakistan of 1.3 beds per 100,000 population. However, such estimates included units which may have no recognised affiliation with critical care training and no support from critical care societies. Whether considering the numbers reported here, or the estimates from those with broader inclusion, CCU bed availability and trained, skilled staff is still lower than neighbouring countries. Pakistan has no central register or standard definition for CCUs and as such units may be operating without the support of trained intensivists.

## Conclusion

This survey provides a detailed landscape of critical care resources and training in Pakistan. Pakistan has an established network of critical care facilities with access to basic equipment but inequalities in access within and between provinces is prominent. Investment in critical care training for doctors and nurses is a key priority for the country. Investment in training for health care staff will likely enable efforts to improve safety within CCUs, accelerate opportunities for research and quality improvement.

## Data Availability

Data is not available on request.

## Ethics approval and consent to participate

A waiver from ethical review was obtained from the National Bioethics Committee of Pakistan.

## Acknowledgments

The authors would like to thank all members of the critical care community in Pakistan who contributed to this work with the shared goal of improving critical care services in the country. We would also like to thank Dr T Tolppa for his support with manuscript preparation, and Mr T Rashan for support with figures.

